# Characterizing Dual Combination Therapy Use in Treatment Escalation of Hypertension: Real-World Evidence from Multinational Cohorts

**DOI:** 10.1101/2021.06.28.21258167

**Authors:** Yuan Lu, Jing Li, Xialin Wang, Jaehyeong Cho, Sreemanee Raaj Dorajoo, Mengling Feng, Min-Huei Hsu, Jason C. Hsu, Jiyoung Hwang, Usman Iqbal, Chen Zhengfeng Jason, Jitendra Jonnagaddala, Yu-Chuan Li, Siaw-Teng Liaw, Hong-Seok Lim, Kee Yuan Ngiam, Phung-Anh Nguyen, Rae Woong Park, Nicole Pratt, Christian Reich, Sang Youl Rhee, Selva Muthu Kumaran Sathappan, Seo Jeong Shin, Hui Xing Tan, Seng Chan You, Xin Zhang, Harlan Krumholz, Marc Suchard, Yun Liu, Mui Van Zandt, Hua Xu

**Author notes:** Listed alphabetically (Jaehyeong Cho, Sreemanee Raaj Dorajoo, Mengling Feng, Min-Huei Hsu, Jason C. Hsu, Jiyoung Hwang, Usman Iqbal, Chen Zhengfeng Jason, Jitendra Jonnagaddala, Yu-Chuan Li, Siaw-Teng Liaw, Hong-Seok Lim, Kee Yuan Ngiam, Phung-Anh Nguyen, Rae Woong Park, Nicole Pratt, Christian Reich, Sang Youl Rhee, Selva Muthu Kumaran Sathappan, Seo Jeong Shin, Hui Xing Tan, Seng Chan You, Xin Zhang).

## Abstract

**Background:** Over one billion adults have hypertension globally, of whom approximately 70% cannot achieve blood pressure control goal with monotherapy alone. Data are lacking on patterns of dual combination therapies prescribed to patients who escalate from monotherapy in routine practice.

**Methods:** Using eleven electronic health record databases that cover 118 million patients across eight countries/regions, we characterized the initiation of antihypertensive dual combination therapies for patients with hypertension. In each database, we first constructed twelve exposure cohorts of patients who newly initiate dual combination therapy with one of the four most commonly used antihypertensive drug classes (angiotensin-converting enzyme inhibitor [ACEi] or angiotensin receptor blocker [ARB]; calcium channel blocker [CCB]; beta-blocker; and thiazide or thiazide-like diuretic) after escalating from monotherapy with one of the three alternative classes. Using these cohorts, we then described dual combination therapy utilization, stratified by age, gender, history of cardiovascular diseases (CVD), and country.

**Results:** Across data sources, we identified 980,648 patients with hypertension initiating dual combination therapy with antihypertensive agents after escalating from monotherapy: 12,541 from Australia, 6,980 from South Korea, 2,096 from Singapore, 7,008 from China, 16,663 from Taiwan, 103,994 from France, 76,082 from Italy, and 754,137 from the United States (US). Significant variations in treatment utilization existed across countries and patient subgroups. In Australia and Singapore, starting an ACEi/ARB monotherapy followed by a CCB was most common while in South Korea, China and Taiwan, starting a CCB monotherapy followed by an ACEi/ARB was most common. In Italy, France, and the US, sequential use of an ACEi/ARB monotherapy followed by a diuretic was most common. Younger patients were more likely to be prescribed ACEi/ARB followed by either a CCB or a diuretic compared with older patients. Women were more likely to be prescribed diuretics then an ACEi/ARB or a CCB compared with men. Among patients with history of CVD, ACEi/ARB followed by beta-blocker, and beta-blocker followed by ACEi/ARB were more commonly prescribed.

**Conclusion:** This is the largest and most comprehensive study characterizing the real-world utilization of dual combination therapies in treating hypertension. Large variation in the transition between monotherapy and dual combination therapy for hypertension was observed across countries. These results highlight the need for future research to identify which second-line dual combination therapy is most effective in practice.

## INTRODUCTION

Hypertension is the leading global risk factor for cardiovascular diseases (CVD) and chronic kidney disease, contributing world-wide to over 7 million deaths and 57 million disability-adjusted life-years annually.^1, 2^ In 2015, approximately 1.13 billion adults had hypertension, yet fewer than 30% have achieved blood pressure (BP) control goal.^3, 4^ Notably, the burden of hypertension is particularly salient in the Asian Pacific region as it has 60% of the world’s population and is experiencing a rapid increase in prevalence of hypertension in the past decades.^3, 5^

Approximately 70% of patients with hypertension cannot achieve BP control goal with monotherapy.^6, 7^ Despite the wide use of combination antihypertensive therapy, considerable uncertainty remains regarding the optimal choice for a second agent with which to escalate from monotherapy. Clinical trials lack head-to-head comparisons of second antihypertensive agents added to monotherapy^8^ and only two trials (ACOMPOLISH and COPE) directly compared different combination regimens in hypertensive patients who require two drugs.^9-11^ These trials, however, provided comparisons between only a few agents, not drug classes, included primarily patients from Western countries, and did not systematically assess heterogeneity in different patient subgroups. The absence of high-quality evidence has limited the ability of clinical guidelines to provide evidence-based recommendations about the preferred choice of the second medication for treatment escalation.^12, 13^ A better understanding of the prescription patterns of dual combination therapy in treatment escalation of hypertension, with attention to relevant subgroups of patients defined by demographic and clinical factors, can lay a foundation for future studies of the comparative effectiveness of different dual antihypertensive combinations.

Accordingly, we conducted a large-scale observational study within the Observational Health Data Science and Informatics (OHDSI) collaborative community to characterize real-world utilization of dual antihypertensive combination therapies for treatment escalation among people with hypertension, using data from eleven electronic health record (EHR) databases across eight countries/regions. We employed a systematic, open science, evidence generation approach for high-quality observational research based on the Large-Scale Evidence Generation and Evaluation across a Network of Databases for Hypertension (LEGEND-HTN) study.^14-16^

## METHODS

### Data Source

We examined patient records from eleven EHR databases mapped to the Observational Medical Outcome Partnership (OMOP) Common Data Model (CDM) version 5.3 from participating research partners across the OHDSI community. These data sources included: IQVIA Longitudinal Patient Database (LPD) Australia (3,101,500 subjects) and Electronic Practice based Research Network 2019 Linked Dataset from South Western Sydney Local Health District (ePBRN SWSLHD, 139,346 subjects) from Australia, Ajou University School of Medicine (AUSOM, 3,109,677 subjects) and Kyung Hee University Hospital (KHMC, 2,010,456 subjects) from South Korea, Khoo Teck Puat Hospital (KTPH, 290,074 subjects) and National University Hospital (NUH, 750,270 subjects) from Singapore, Jiangsu Province Hospital (6,230,000 subjects) from China, Taipei Medical University Clinical Research Database (TMUCRD) (3,659,572 subjects) from Taiwan, IQVIA LPD France (18,118,000 subjects) from France, IQVIA LPD Italy (2,209,600 subjects) from Italy, and IQVIA US Ambulatory EMR (78,526,000 subjects) from the United States (US, **Appendix Table 1**). These data partners altogether monitor over 118 million patients from eight countries and regions across the world.

We executed this study through the federated network model of OHDSI, where access to data and statistical analyses were executed inside each data partner’s institution using the OHDSI common toolstack.^14, 15, 17-19^ We prespecified the entire analytical process before execution and collected aggregated results from data partners for interpretation. Each data partner obtained the necessary Institutional Review Board (IRB) approval or exemption. Previous studies have demonstrated that this process could be successfully applied to evaluate the comparative effectiveness of first-line antihypertensive monotherapies.^16, 20, 21^

### Study Population

The study population consisted of adult patients (≥18 years) with prior antihypertensive monotherapy who newly initiate escalated treatment with one of the 56 drug ingredients (**Appendix Table 2**) that comprise four major drug classes from 2000 to 2019. For these patients, we constructed 12 non-overlapping, exposure cohorts. Each cohort contains new-users of one of the four dual combination drug classes after escalating from monotherapy with one of the three alternative classes (**Appendix Table 3**).

New-user cohort design is advocated as the primary design choice for comparative effectiveness research.^22-24^ By identifying patients who start a new drug for treatment escalation and using initiation of the second drug as the start of follow-up, the new-user design models a randomized controlled trial (RCT) where the randomized intervention commences upon treatment escalation at the patient’s index visit.

Specifically, cohort entry (index date) for each patient was their date of prescription initiating the second drug containing the RxNorm ingredient concept codes of the four major drug classes from 2000 to 2019. Inclusion criteria for patients based on the index date included: 1) at least one hypertension diagnosis any time in the patient’s record before the index date; 2) at least one year of observation time before the index date (washout period to improve new-user sensitivity); 3) at least one prescription of an antihypertensive agent and no prescriptions for any other agent any time before the index date; 4) at least 30 days between the initiation of first drug class and the initiation of second drug class on the index date (**Appendix Figure 1**). We purposefully did not exclude patients with a history of cardiovascular events (CVD), enabling us to report drug utilizations for individuals with and without history of CVD. History of CVD was defined by at least one diagnosis code for arteriosclerotic vascular disease, cerebrovascular disease, ischemic heart disease, or peripheral vascular disease any time on or prior to the index date. Continuous drug exposures were constructed by allowing fewer than 30-day gaps between prescriptions.

**Figure 1.**
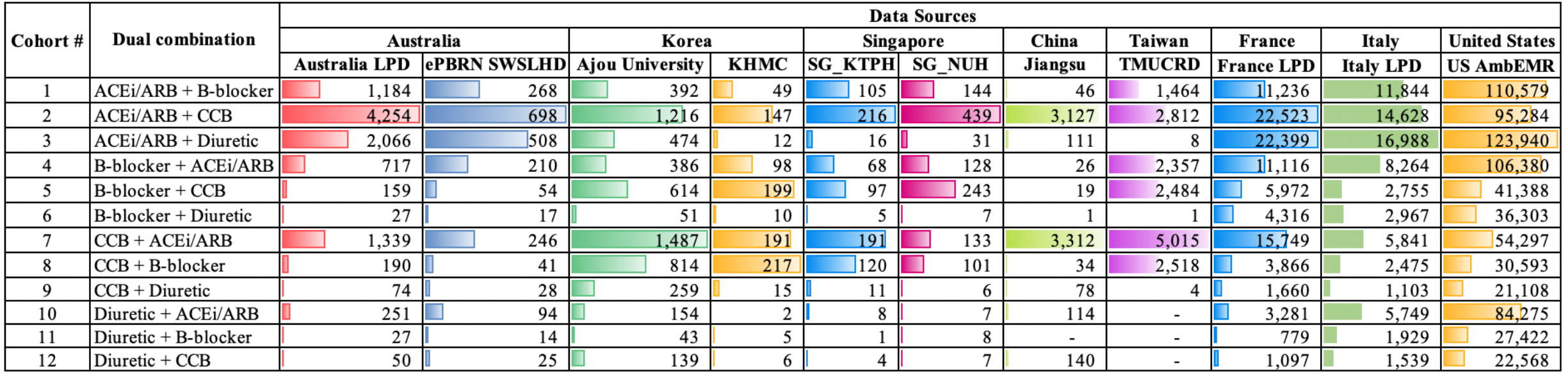
Patient counts of 12 exposure cohorts from 11 committed data sources*. *ACEi/ARB + B-Blocker denotes starting an ACEi/ARB monotherapy followed by a beta-blocker. ACEi = angiotensin converting enzyme inhibitor; ARB = angiotensin converting enzyme inhibitor; B-blocker = beta-blocker; CCB = calcium channel blocker.

### Cohort Development and Validation

We developed all exposure cohorts above using OHDSI’s open source ATLAS^25^ platform that enables researchers to define cohorts based on drug exposures, diagnoses, procedures and patient characteristics through a user-friendly interface. We based drug exposure on occurrences of RxNorm codes in the appropriate OMOP CDM tables and built diagnosis concept sets, such as “hypertension diagnosis,” as SNOMED term collections in the appropriate OMOP CDM tables. ATLAS enforced complete transparency in cohort definitions by automatically generating human-and computer-readable representations. We used previously validated concept definitions for hypertension diagnosis and antihypertensive agents.^16^

We further validated exposure cohorts and aggregated drug utilization using comprehensive cohort characterization tools against data sources through the OHDSI’s CohortDiagnostic package.^26^ For each cohort and data source, this package systematically generated incidence new-user rates (stratified by age, gender, and calendar year), cohort characteristics (all comorbidities, drug use, health utilization) and the actual codes found in the patient records triggering the various rules in the cohort definitions. This approach allowed us to better understand the heterogeneity of source coding for exposures and health outcomes as well as the impact of various inclusion criteria on overall cohort counts.

### Statistical Analysis

For each database, we described the overall utilization in dual combination therapies and evaluated treatment variation in patient groups by age (18-44 / 45-64 / ≥65 years), gender, history of CVD, and country. Specifically, we calculated the proportion of users of each dual combination regiment. We compared the distribution of treatment utilization between patient subgroup defined by age, gender, and history of CVD using Chi-squared tests or t-tests. A pre-specified 2-sided p-value of < 0.05 will be used to indicate statistical significance. Finally, we characterized treatment pathways for hypertension (i.e., the ordered sequence of medications that a patient is prescribed) in diverse populations using the Sunburst plots. The sequences included changes in medication and additions of medication.

## RESULTS

### Utilization of dual combination therapies in treatment escalation

Among 118 million patients identified across eleven data sources, we found 980,648 patients with hypertension being newly initiating dual combinations of antihypertensive agents after escalating from monotherapy: 12,541 from Australia, 6,980 from South Korea, 2,096 from Singapore, 7,008 from China, 16,663 from Taiwan, 103,994 from France, 76,082 from Italy, and 754,137 from the US (**Figure 1**). Patient ages varied across data sources, but most patients clustered around the ages of 45 to 64 years old. The proportion of women was similar as that of men.

We observed significant variation in treatment utilization across countries. Starting an ACEi/ARB monotherapy followed by a CCB was the most commonly prescribed combination in Australia (proportion of users ranged from 45.2-45.9% across databases) and Singapore (12.9-23.2%), while starting a CCB monotherapy followed by an ACEi/ARB was most common in South Korea (19.7-23.3%) and China (38.3%). Sequential use of an ACEi/ARB monotherapy followed by a diuretic was most common in Italy (28.5%), France (27.4%), and the US (25.2%). ACEi/ARB monotherapy followed by a beta-blocker, or beta-blocker monotherapy followed by an ACEi/ARB were more commonly prescribed in the US and Italy than in other countries. CCB monotherapy followed by a beta-blocker, or beta-blocker monotherapy followed by a CCB were more commonly prescribed in the South Korea. Certain drug combinations, including CCB or beta-blocker monotherapy followed by a diuretic, or diuretic monotherapy followed by a CCB or a beta-blocker were among the least frequently prescribed combinations across all countries (proportion of users ranged from 0.6% to 1.2%).

Figure 2 shows the proportion of users of each dual combination regiment by age and sex. Most (33-57%) of the patients who were starting an ACEi/ARB monotherapy followed by a CCB or starting an ACEi/ARB followed by a diuretic were between 45 and 64 years of age, whereas most (28-73%) of those who were starting an CCB followed by a diuretic, starting a CCB followed by a beta-blocker, or starting an ACEi/ARB followed by a beta-Blocker were in age of 65 years or older. Compared with men, women were more likely to be prescribed a diuretic monotherapy followed by an ACEi/ARB (38-74%) or a beta-blocker followed by a diuretic (19-73%). Men were more likely to be prescribed am ACEI/ARB monotherapy followed by a beta-blocker (47-63%), or a beta-blocker followed by an ACEi/ARB (39-58%). **Figure 3** reports the proportion of users of each dual combination regiment by history of CVD. Over 80% of the hypertensive patients included in our analysis did not have history of CVD. Among patients with history of CVD, starting an ACEi/ARB monotherapy followed by a beta-blocker, or starting a beta-blocker followed by an ACEi/ARB were the most commonly prescribed dual combinations.

### Treatment pathway for hypertension

Tracking medication changes for these 980,648 patients over time revealed a diverse array of treatment trajectories across countries. **Figure 4** illustrates the treatment pathways for hypertensive agents across the largest nine data sources. The most common first-line therapy of patients in Australia and Singapore was an ACEi/ARB, whereas the most common first-line therapy of patients in South Korea was a CCB. The proportion of patients who were prescribed with dual therapy differed between countries. There were more patients in Australia who were initiated with dual therapy than patients in South Korea. Most patients (84.1%) in China initiated with a CCB or an ACEi/ARB. The commonly prescribed agents in the US, Italy and France were an ACEi/ARB and a diuretic.

**Figure 2.**
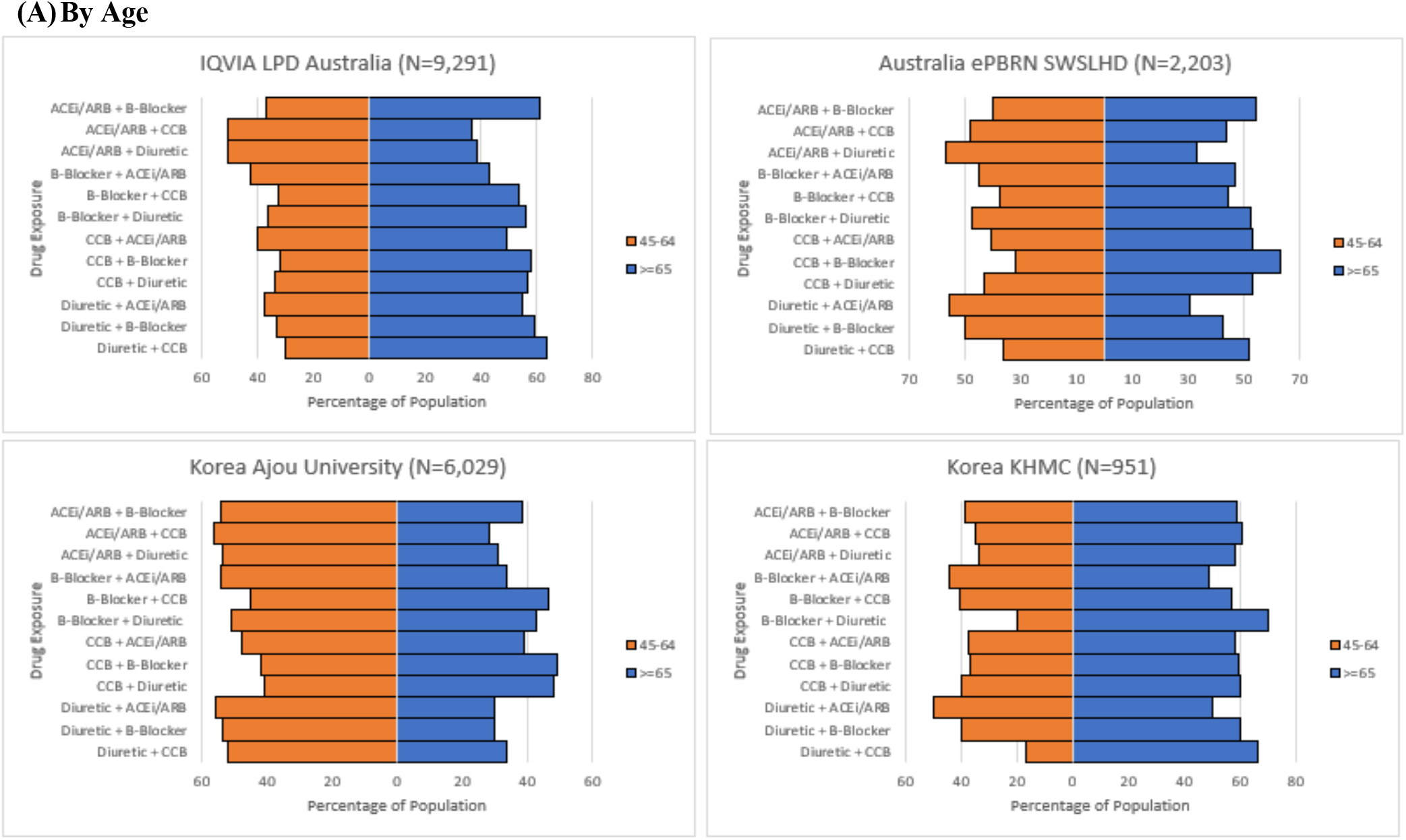
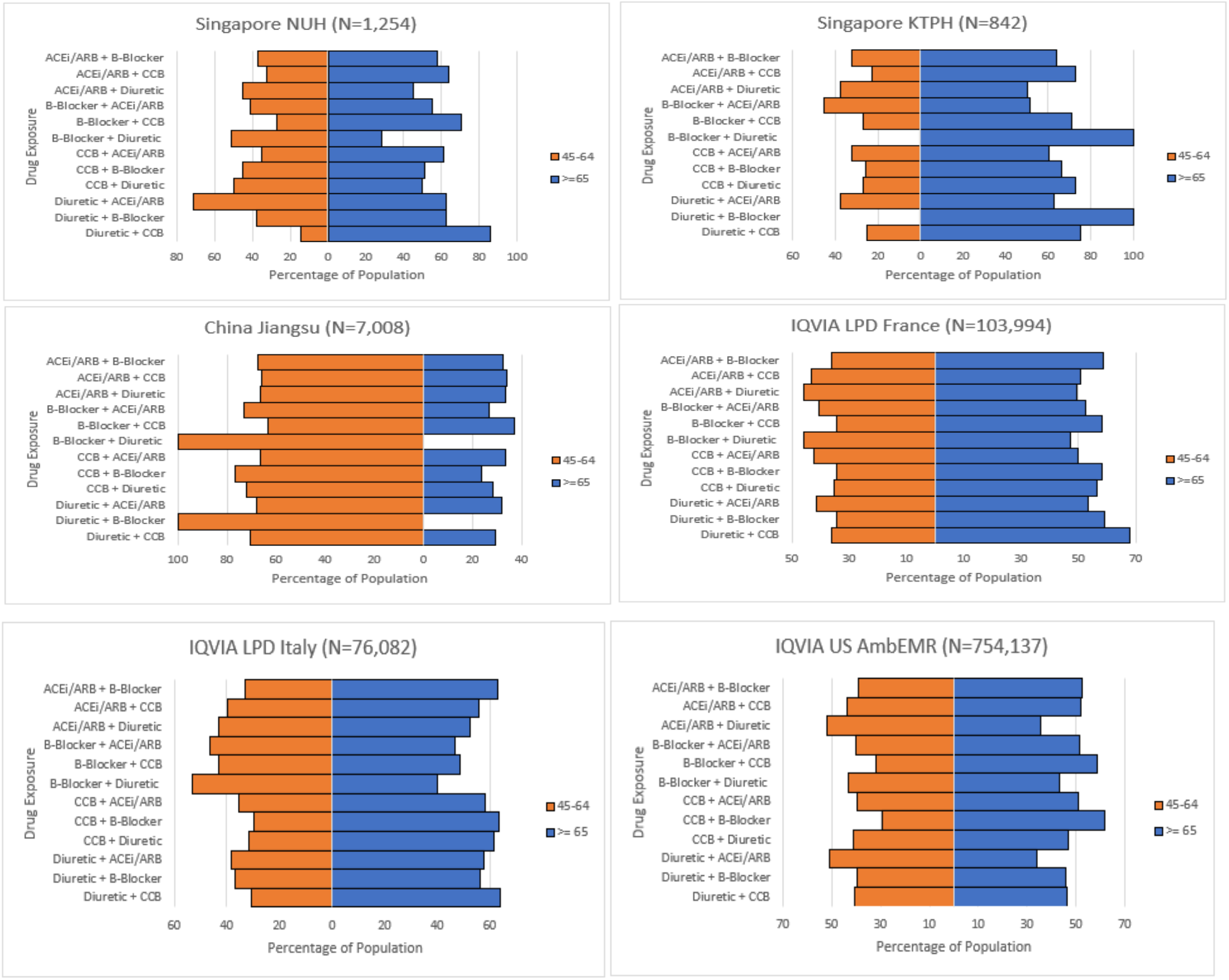
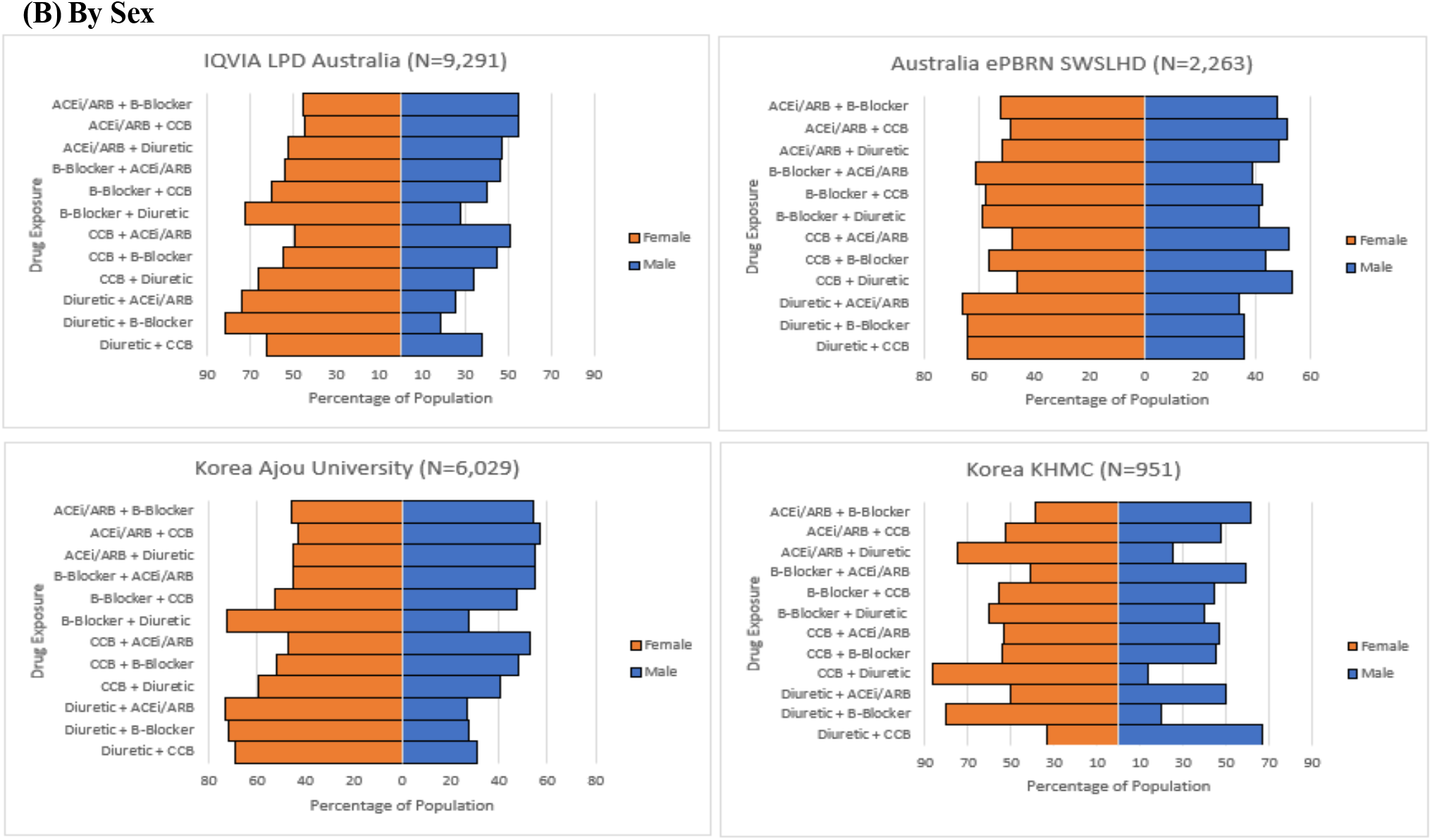
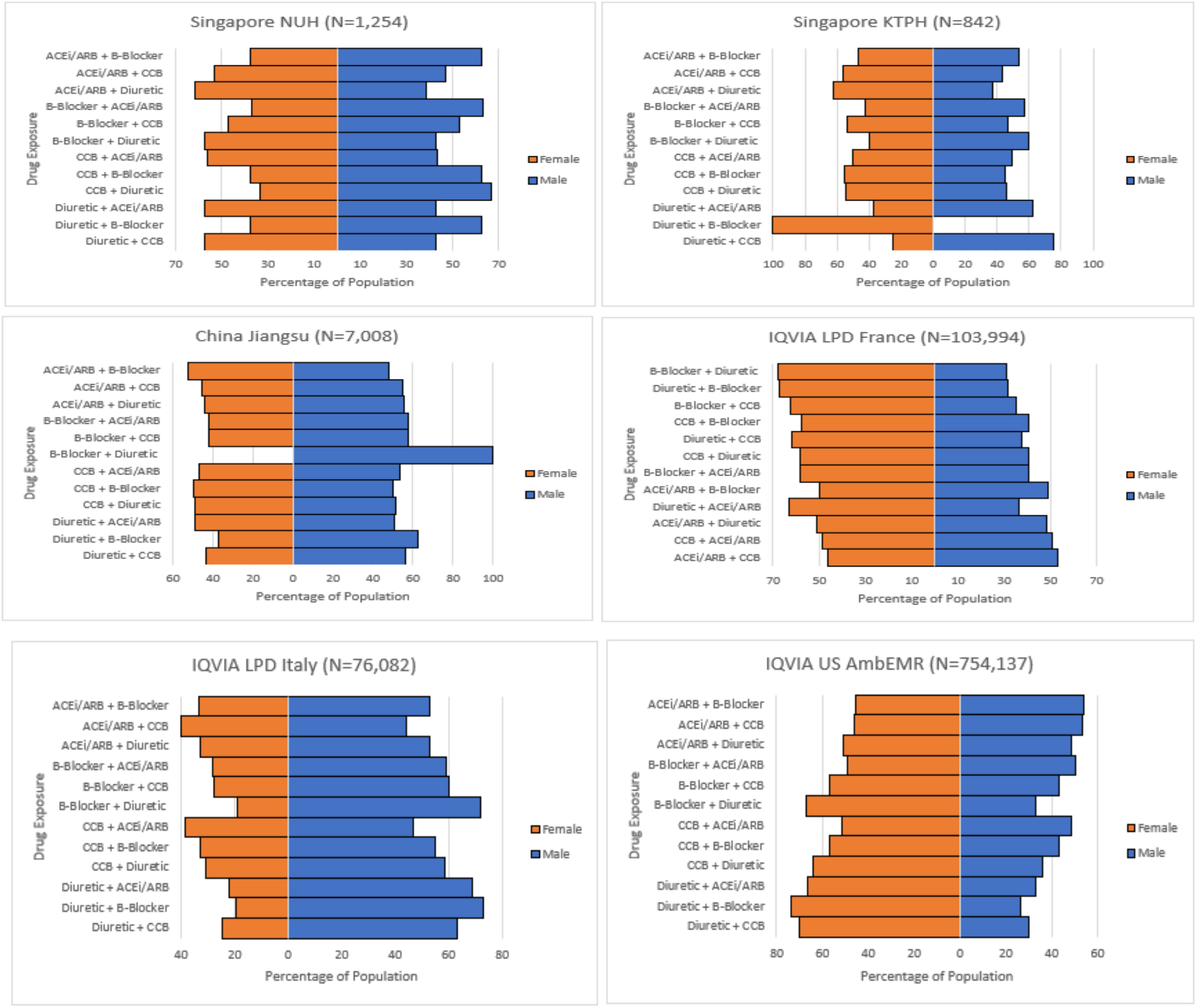
Proportion of users of each dual combination of antihypertensive agents by (A) age and (B) gender*. *ACEi/ARB + B-Blocker denotes starting an ACEi/ARB monotherapy followed by a beta-blocker.

**Figure 3.**
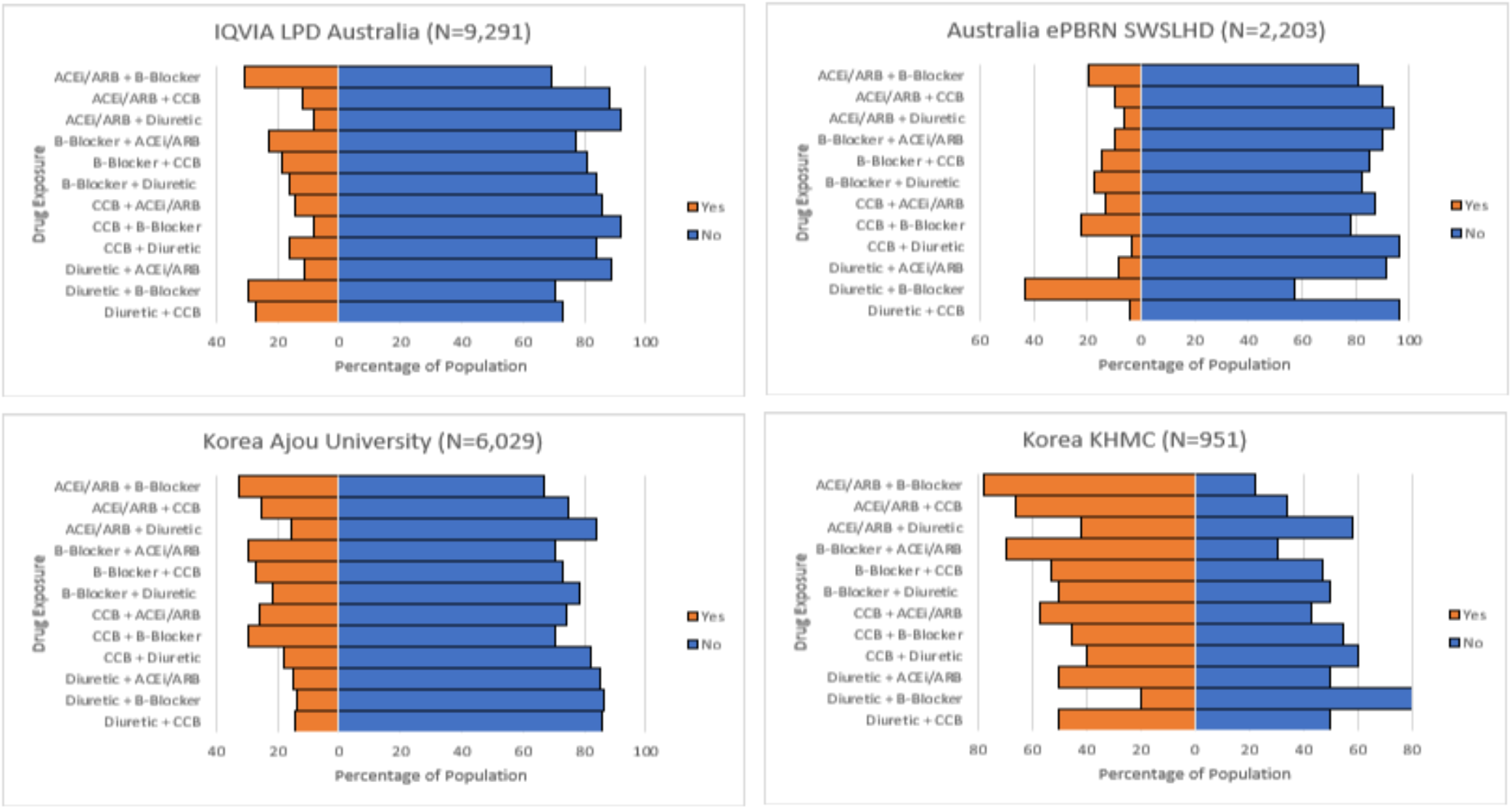
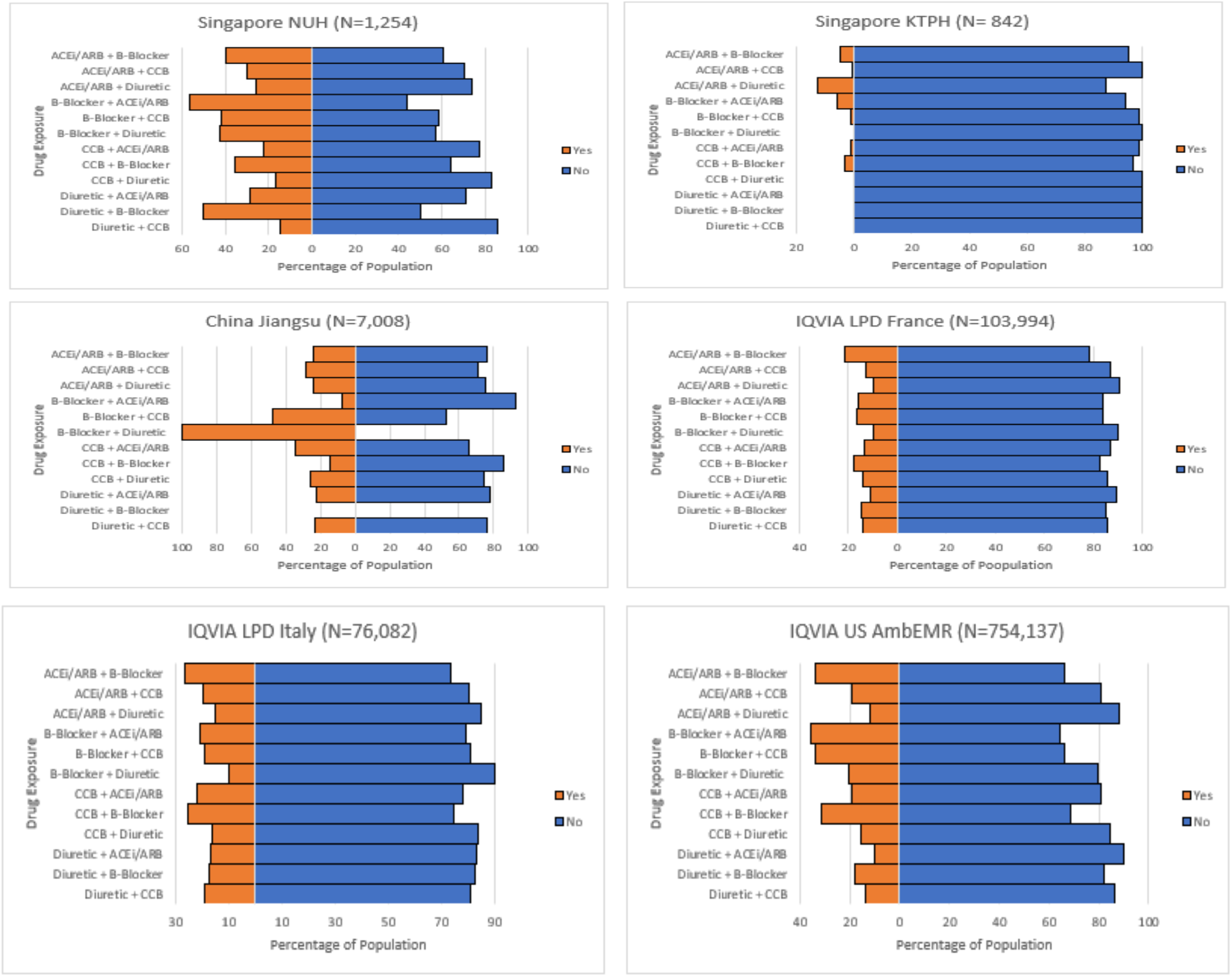
Proportion of users of each dual combination of antihypertensive agents by history of cardiovascular diseases.

**Figure 4.**
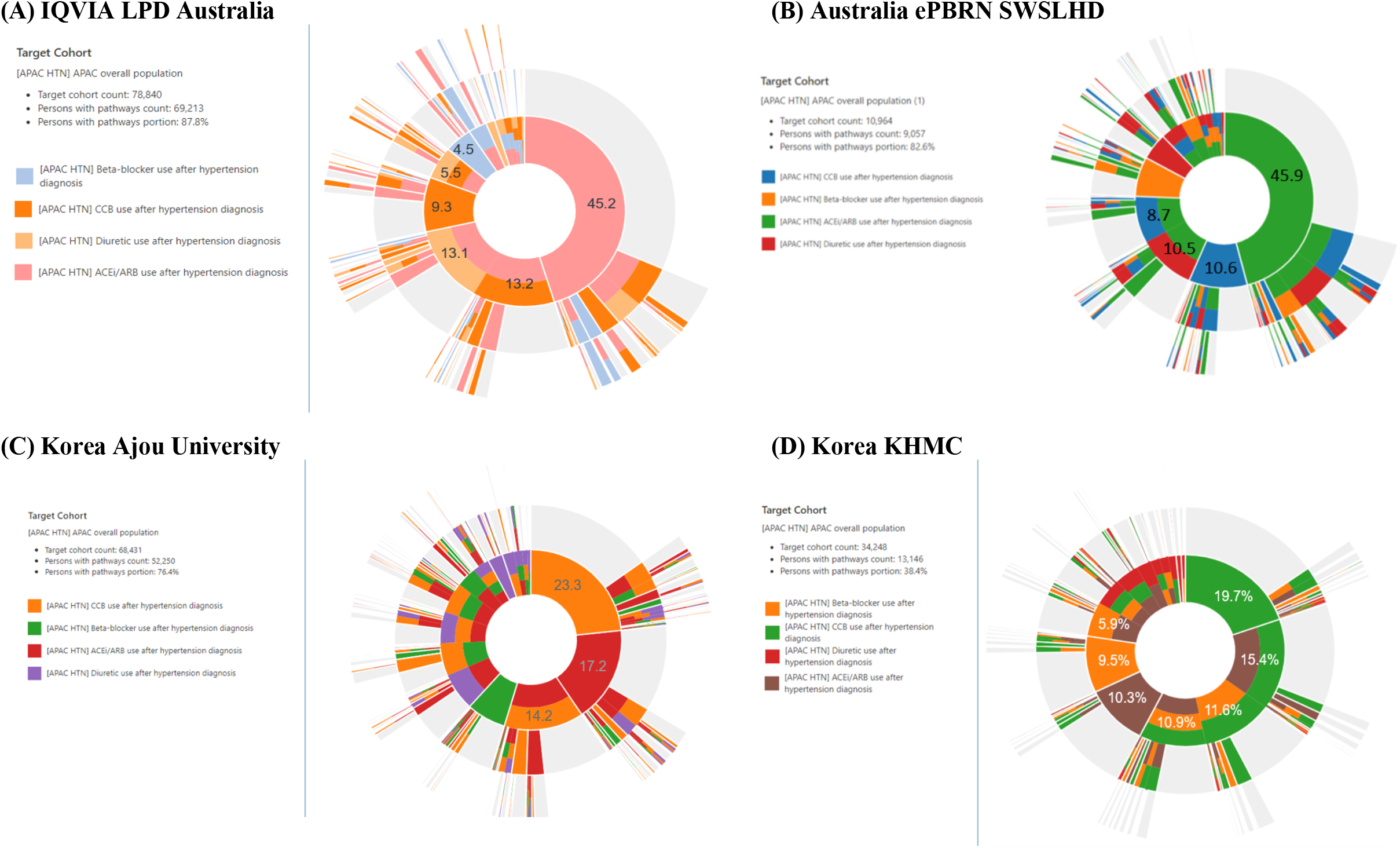
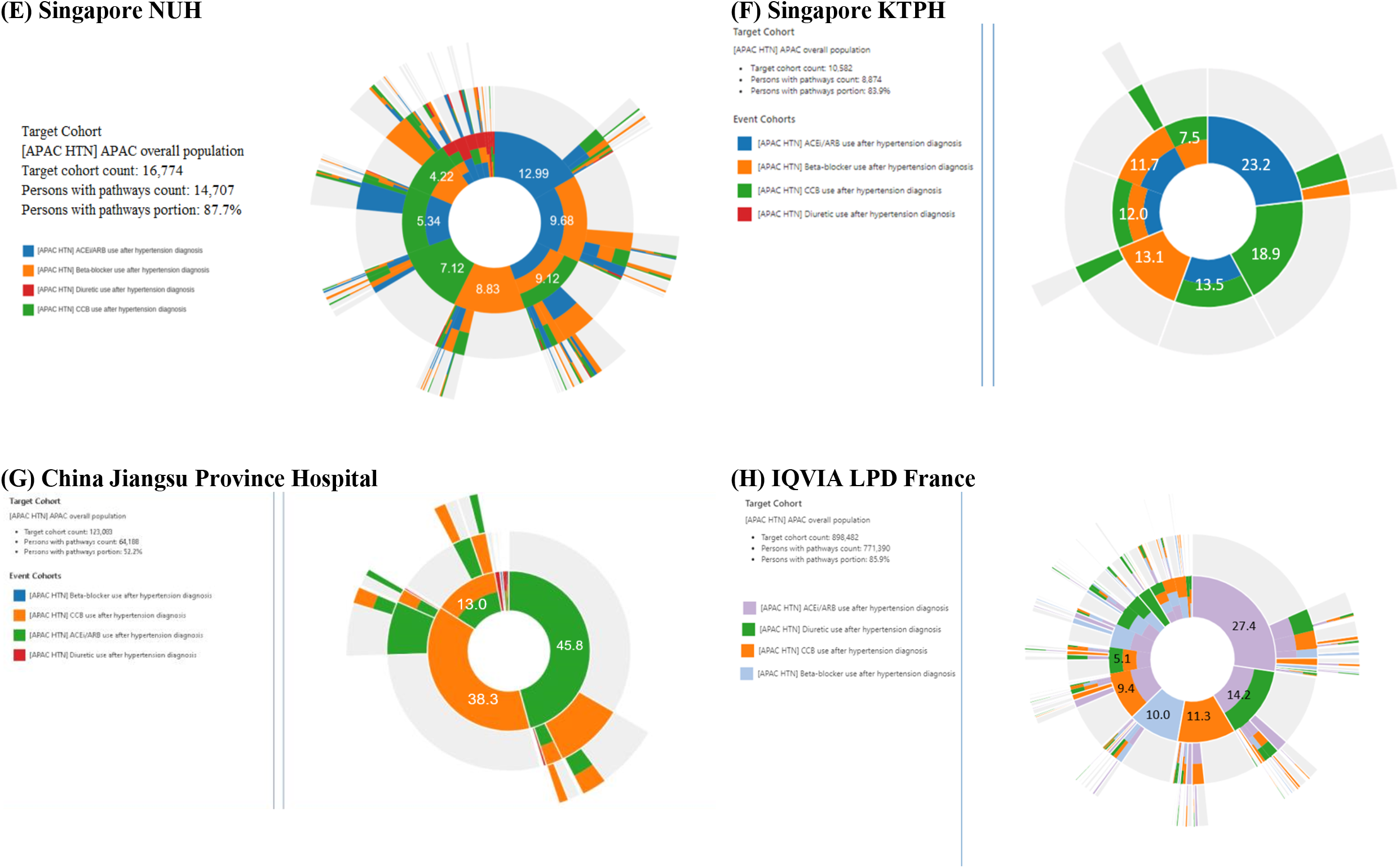
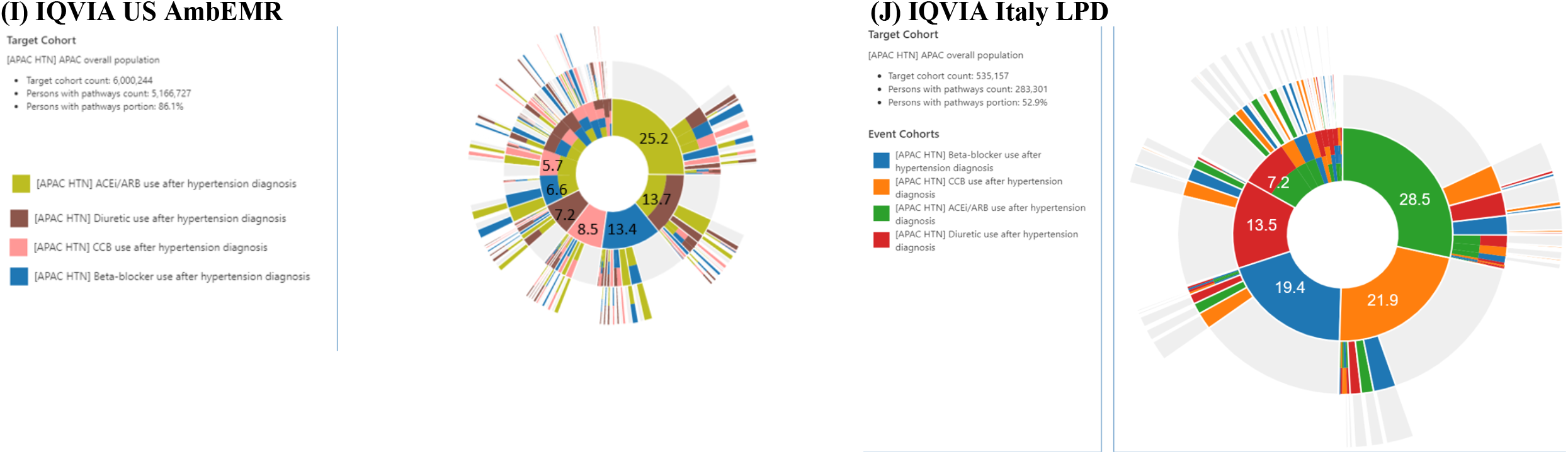
Treatment pathway of hypertension.

## DISCUSSION

This is the first study to describe the real-world utilization of antihypertensive dual combination therapies for treatment escalation across eight countries, including five Asia Pacific countries and regions. We observed heterogeneity in the use of dual combination therapies as recorded in EHR and administrative claim data sources, identifying a total of 980,648 patients with hypertension and dual combination therapy in Australia, South Korea, Singapore, China, Taiwan, France, Italy and the United States. These findings provide insight into the current prescription patterns of antihypertensive agents and are critical to guide the real-world application of treatment decision pathways for patients with hypertension.

Our study extends the prior literature substantially as it is the largest multisite analysis of real-world evidence to address dual combination therapies used in treatment escalation of hypertension. Through the OHDSI network (particularly the OHDSI APAC network), we take advantage of disparate health databases drawn from different sources and across a range of countries and practice settings. These large-scale and unfiltered populations better represent real-world practice than the restricted study populations from RCTs and population-based cohort studies. This first descriptive analysis of the OHDSI APAC collaborative demonstrates that coordinated efforts can overcome many of the logistic and methodological challenges associated with observational study designs. The profiles of treatment pathways are based on more than 118 million patient records. We successfully addressed patient privacy and diverse research regulatory constraints, adopted a consistent data model, and distributed queries across a broad population.

There are several possible explanations to our findings. The observed prescription pattern of antihypertensive agents is, in part, a reflection of hypertension guidelines issued in the past few decades. An ACEi/ARB is the most commonly prescribed drug class across all data sources as it is recommended as a first-line treatment option by most guidelines.^12, 27-31^ The finding of CCBs being the predominant prescribed drug class in the Chinese data source is consistent with a previous national study in China,^32^ which may reflect the endorsement of the clinical guideline in China and the lower cost of CCBs compared with other antihypertensive drugs.^31^ Despite beta-blockers being less effective,^33, 34^ their high use in South Korea and US is consistent with nationwide studies in those countries that revealed the use of beta-blocker monotherapy for hypertensive patients remains prevalent.^16, 29, 35^ Among patients with a history of CVD, the common use of an ACEi/ARB or a beta-blocker stands consistent with guidelines for secondary prevention of CVD.^36-38^ Finally, our study corroborates the previous work by OHDSI researchers, which revealed significant heterogeneity in treatment pathways for several chronic diseases across data sources.^16, 20, 39^

Our findings also have important public health implications. The heterogeneity of treatment pathways of hypertension across countries and sites reflects the failure of the field to converge on effective consistent treatment escalation algorithm for hypertension. Current guidelines do not provide recommendations about the preferred choice of the second agent added to monotherapy due to the lack of evidence from RCT,^12, 27-31^ and the large variation observed in clinical practice may reflect a trial-and-error approach to intensify treatment for hypertension. This finding highlights the need to generate robust real-world evidence on the efficacy and safety of different combinations of antihypertensive agents. While RCTs remain a key tool for high-quality clinical efficacy estimates in controlled settings, real-world observational studies can help to fill evidence gaps where large-scale RCTs are not feasible, such as in the study of second-and third-line hypertension treatments.

Several limitations need to be considered when interpretating the results of this study. First, our study was based on routinely collected real-world data, where misclassification of diseases and therapies may be present. We only included patients who had a clinical diagnosis of hypertension; therefore, patients without a coded diagnosis would have been excluded even if they had elevated blood pressure that met the criteria of hypertension. Second, treatment misclassification is possible given that participating data sources varied in their capture of drugs, from hospital billing records, prescription orders, or dispensing data. Third, the lack of information on medication is another limitation as this is important information that may add value to the understanding of prescribing trends. Fourth, this study only describes prescription pattern of antihypertensive medications and we do not have information about medication compliance among patients with hypertension. Finally, our study is limited to eight countries and regions, of which the findings may not be generalizable to other countries of the world.

In conclusion, this is the largest and most diverse study characterizing the real-world utilizations of dual combination therapies in treatment escalation of hypertension. Large variation in drug utilization was observed in routine practice, highlighting the need for future research on the safety and efficacy of the more commonly used treatments.

## Data Availability

All aggregated data and executable source code are available through GitHub at: github.com/ohdsi-studies.

## Acknowledgement

The authors appreciate Patrick Ryan and George Hripcsak for their inputs into this paper.

## Ethical approval

All the data partners received Institutional Review Board (IRB) approval or exemption.

| Data Source | IRB Approval |
| --- | --- |
| IQVIA LPD Australia | Use of de-identified IQVIA data sources was deemed not human subject research by the IQVIA internal review committee and approved for OHDSI network studies. |
| ePBRN SWSLHD 2019 Linked Dataset | Use of Australia ePBRN SWSLHD data source was approved by the UNSW Sydney and South Western Sydney Local Health District Human Research Ethics Committees (Project number: 2019/PID05368). |
| Korea Ajou University School of Medicine (AUSOM) CDM | The Korean Bioethics Act does not require an IRB review for retrospective observational studies using common data model in distributed research networks. |
| Korea CDM of Kyung Hee University Hospital | The Korean Bioethics Act does not require an IRB review for retrospective observational studies using common data model in distributed research networks. |
| Khoo Teck Puat Hospital (KTPH) | Use of KTPH data source was reviewed by National Health Group Domain Specific Review Board and approved the request for waiver of informed consent (Project number: 2017/00995). |
| National University Hospital (NUH) | Use of NUH data source was reviewed by National Health Group Domain Specific Review Board and was determined that it qualifies for exemption because the analysis involved a dataset without identifiers (Project number: 2021/00125). |
| China Jiangsu Province Hospital | Use of Jiangsu Province Hospital data source was reviewed and approved by the Nanjing Medical University Institutional Review Board. |
| Taiwan Taipei Medical University Clinical Research Database (TMUCRD) | Use of TMUCRD data has been approved by the TMU-Joint Institutional Review Board (Project number: TMU-JIRB N202011003). |
| IQVIA Ambulatory EMR | Use of de-identified IQVIA data sources was deemed not human subject research by the IQVIA internal review committee and approved for OHDSI network studies. |
| IQVIA LPD France | Use of de-identified IQVIA data sources was deemed not human subject research by the IQVIA internal review committee and approved for OHDSI network studies. |
| IQVIA LPD Italy | Use of de-identified IQVIA data sources was deemed not human subject research by the IQVIA internal review committee and approved for OHDSI network studies. |

