## Supplemental material for "Characterizing Dual Combination Therapy Use in Treatment Escalation of Hypertension: Real-World Evidence from Multinational Cohorts"

Supplementary Online Content


| **Title** | **Page** |
|---|---|
| Appendix Text. Description of IQVIA data sources | 2 |
| Appendix Table 1. Data Source Description | 3 |
| Appendix Table 2. Drug codes of 56 drug ingredients in four major antihypertensive drug classes | 4 |
| Appendix Table 3. Twelve exposure cohorts for Class vs. Class comparison | 5 |
| Appendix Figure 1. Graphical presentation of cohort definitions | 6 |

**Appendix Text. Description of IQVIA data sources**

**IQVIA LPD Australia**
The IQVIA LPD Australia database consists of anonymized patient records collected from Patient Management software used by GPs during an office visit to document patients' clinical records. The data originates from two sources: longitudinal patient data and practice profiles where they are integrated into one common data source.

**IQVIA US Ambulatory EMR**
The IQVIA US Ambulatory EMR database consists of longitudinal, de-identified electronic health records originating from ambulatory clients. The data contains detailed clinical information that captures important health outcomes such as lab test results and vital signs. It also covers administered drugs including prescription and over-the-counter medicines, vaccines, large-molecule biologic therapies, route of administration, days suppl and refill information.

**IQVIA Longitudinal Patient Database (LPD) France**
The IQVIA OMOP Longitudinal Patient Database (LPD) France database consists of anonymized patient records collected from Patient Management software used by Doctors during an office visit to document patients' clinical records. The total database consists of 1200 GPs, 7.8 million patients, 620 specialists across 8 specialties (cardiology, neurology, psychiatry, pulmonology, gastroenterology, gynecology, diabetology & rheumatology)

**IQVIA LPD Italy**
LPD Italy is comprised of anonymized patient records collected from software used by GPs during an office visit to document patients' clinical records. Data coverage includes over 2M patient records with at least one visit and 119.5M prescription orders across 900 GP practices. Dates of service include from 2004 through present. Observation time is defined by the first and last consultation dates. Drugs are captured as prescription records with product, quantity, dosing directions, strength, indication and date of consultation.

**Appendix Table 1. Data Source Description**

| Data Source | Data Type | Country/District | Time Period | No. of Patients |
|---|---|---|---|---|
| IQVIA LPD Australia | EHR | Australia | 2006-2020 | 3,101,500 |
| ePBRN SWSLHD 2019 Linked Dataset | EHR | South Western Sydney, Australia | 2012-2019 | 139,346 |
| Korea Ajou University School of Medicine (AUSOM) CDM | EHR | Suwon, Korea | 1995-2019 | 3,109,677 |
| Korea CDM of Kyung Hee University Hospital | EHR | Seoul, Korea | 2008-2018 | 2,010,456 |
| Khoo Teck Puat Hospital (KTPH) | EHR | Singapore | 2010-2016 | 290,074 |
| National University Hospital (NUH) | EHR | Singapore | 2015-2018 | 750,270 |
| China Jiangsu Province Hospital | EHR | China | 2005-2015 | 6,230,000 |
| Taiwan Taipei Medical University Clinical Research Database (TMUCRD) | EHR | Taiwan | 2004-2020 | 3,659,572 |
| IQVIA Ambulatory EMR | EHR | United States | 2006-2020 | 78,526,000 |
| IQVIA LPD France | EHR | France | 1994-2020 | 18,118,000 |
| IQVIA LPD Italy | EHR | Italy | 2004-2020 | 2,209,600 |

*EHR = Electronic health record

### Appendix Table 2. Drug codes of 56 drug ingredients in four major antihypertensive drug classes

| ACEI/ARB | | | Beta-blocker | | | Calcium channel blocker | | | Thiazide Diuretics | | |
|---|---|---|---|---|---|---|---|---|---|---|---|
| Ingredient name | RxNorm | OMOP ID | Ingredient name | RxNorm | OMOP ID | Ingredient name | RxNorm | OMOP ID | Ingredient name | RxNorm | OMOP ID |
| Benazepril | 18867 | 1335471 | Esmolol | 49737 | 19063575 | Diltiazem | 3443 | 1328165 | Hydrochlorothiazide | 5487 | 974166 |
| Captopril | 1998 | 1340128 | Celiprolol | 20498 | 19049145 | Verapamil | 11170 | 1307863 | Xipamide | 11371 | 19010493 |
| Cilazapril | 21102 | 19050216 | Oxprenolol | 7801 | 19024904 | Amlodipine | 17767 | 1332418 | Chlorthalidone | 2409 | 1395058 |
| Enalapril | 3827 | 1341927 | Labetalol | 6185 | 1386957 | Nifedipine | 7417 | 1318853 | Indapamide | 5764 | 978555 |
| Fosinopril | 50166 | 1363749 | Propranolol | 8787 | 1353766 | Nicardipine | 7396 | 1318137 | Metolazone | 6916 | 907013 |
| Imidapril | 60245 | 19122327 | Carvedilol | 20352 | 1346823 | Felodipine | 4316 | 1353776 | | | |
| Lisinopril | 29046 | 1308216 | Pindolol | 8332 | 1345858 | Nisoldipine | 7435 | 1319880 | | | |
| Moexipril | 30131 | 1310756 | Bisoprolol | 19484 | 1338005 | Isradipine | 33910 | 1326012 | | | |
| Perindopril | 54552 | 1373225 | Penbutolol | 7973 | 1327978 | Nimodipine | 7426 | 1319133 | | | |
| Quinapril | 35208 | 1331235 | Betaxolol | 1520 | 1322081 | Clevidipine | 233603 | 19089969 | | | |
| Ramipril | 35296 | 1334456 | Acebutolol | 149 | 1319998 | Nilvadipine | 53692 | 19113063 | | | |
| Trandolapril | 38454 | 1342439 | Nebivolol | 31555 | 1314577 | Mepirodipine | 39879 | 19102106 | | | |
| Zofenopril | 39990 | 19102107 | Atenolol | 1202 | 1314002 | Manidipine | 29275 | 19071995 | | | |
| Valsartan | 69749 | 1308842 | Metoprolol | 6918 | 1307046 | Nitrendipine | 7441 | 19020061 | | | |
| Candesartan | 214354 | 1351557 | Carteolol | 2116 | 950370 | Lercanidipine | 135056 | 19015802 | | | |
| Eprosartan | 83515 | 1346686 | | | | Lacidipine | 28382 | 19004539 | | | |
| Irbesartan | 83818 | 1347384 | | | | | | | | | |
| Losartan | 52175 | 1367500 | | | | | | | | | |
| Olmesartan | 321064 | 40226742 | | | | | | | | | |
| Telmisartan | 73494 | 1317640 | | | | | | | | | |

OMOP ID indicates the exact code used in OMOP-CDM based on RxNorm ontology. Corresponding RxNorm code to OMOP-ID are also described in this table.
ACEI = angiotensin converting enzyme inhibitor; ARB = angiotensin converting enzyme inhibitor; CDM = common data model; OMOP = observational medical outcomes partnership.

**Appendix Table 3. Twelve exposure cohorts for Class vs. Class comparison**

| Cohort # | Dual combination* |
|---|---|
| 1 | ACEi/ARB + B-blocker |
| 2 | ACEi/ARB + CCB |
| 3 | ACEi/ARB + Diuretic |
| 4 | B-blocker + ACEi/ARB |
| 5 | B-blocker + CCB |
| 6 | B-blocker + Diuretic |
| 7 | CCB + ACEi/ARB |
| 8 | CCB + B-blocker |
| 9 | CCB + Diuretic |
| 10 | Diuretic + ACEi/ARB |
| 11 | Diuretic + B-blocker |
| 12 | Diuretic + CCB |

* ACEi/ARB + B-Blocker denotes starting an ACEi/ARB monotherapy followed by a beta-blocker.

ACEi = angiotensin converting enzyme inhibitor; ARB = angiotensin converting enzyme inhibitor; B-blocker = beta- blocker; CCB = calcium channel blocker.

**Appendix Figure 1. Graphical presentation of cohort definitions**

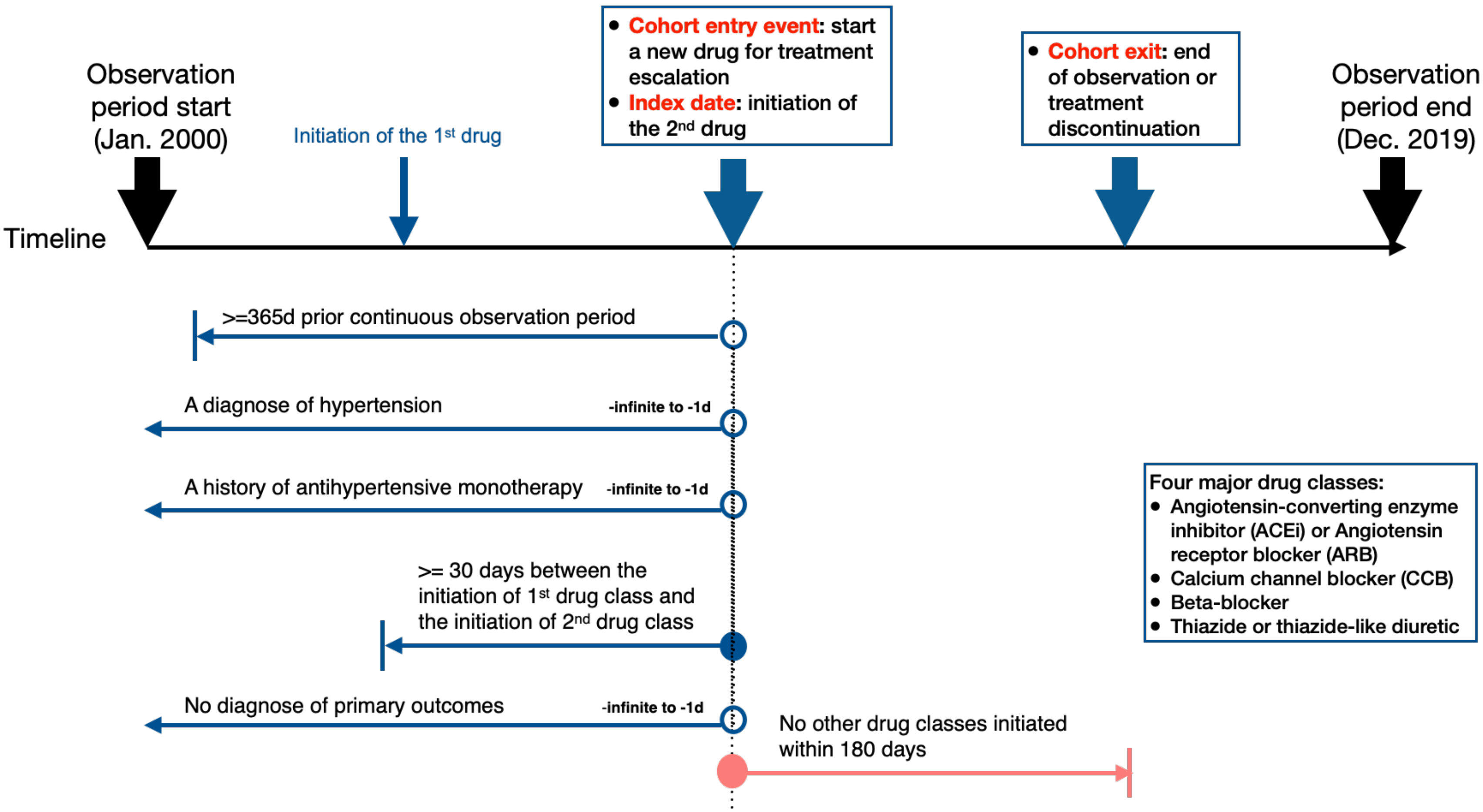
